# MLX-Radiomics: accelerating standardized regional and voxel-wise radiomics for CT, PET and MRI

**DOI:** 10.64898/2026.11.08.26367560

**Authors:** Joseph Sandoval, Alfredo Sandoval, Arsalan Saleem

**Author notes:** Corresponding authors: Alfredo Sandoval Jr.,; Arsalan Saleem. These authors contributed equally (Joseph Sandoval, Alfredo Sandoval).

## Abstract

**Background:** Differences among radiomics implementations complicate cross-study comparisons, while voxel-wise feature extraction remains computationally demanding. We developed MLX-Radiomics to accelerate regional feature extraction and voxel-wise mapping while preserving agreement with standardized reference measurements.

**Methods:** MLX-Radiomics implements 172 feature definitions from the Image Biomarker Standardization Initiative (IBSI), 16 extensions and the IBSI 2 image filters. We assessed agreement with IBSI reference data and evaluated numerical accuracy against independent double-precision and analytic calculations. In matched comparisons on Apple M1 Max and M5 Max systems, we used the IBSI reference CT to compare feature values, extraction time and memory use with PyRadiomics, MIRP and Flash-Radiomics, and feature values with FastRad. Separately, we compared the time required to generate voxel-wise feature maps for regions of up to 10.1 million voxels with published GPU results obtained on different hardware and images. We evaluated agreement with MIRP under published settings in a 51-patient cohort with CT, FDG PET and T1-weighted MRI.

**Results:** MLX-Radiomics agreed with all 2,160 IBSI reference measurements within the specified tolerances. With matching outputs, MLX-Radiomics was 2.66 to 7.27 times faster than PyRadiomics and MIRP for regional feature extraction and 198 to 218 times faster than PyRadiomics for voxel-wise mapping on a 24^3^-voxel crop. It also outperformed the fastest Flash-Radiomics CPU configurations tested. On the M5 Max, generating 93 maps over 10.1 million voxels took 23.8 s. In the patient cohort, MLX-Radiomics and MIRP agreed on 98.0% of requested values. For tumors at the image edge, matching the libraries’ resampling rules substantially improved agreement.

**Conclusions:** MLX-Radiomics provides standardized regional measurements and voxel-wise maps for CT, PET and MRI, with evidence of faster extraction limited to CT. It can support quantitative imaging research on relatively inexpensive hardware without requiring a separate GPU workstation.

## Introduction

Radiomic measurements are influenced by scanner and acquisition parameters, reconstruction, and software implementation [1], [2]. Even with identical images and segmentations, differences in preprocessing and feature definitions can yield different measurements [2]. This variability complicates comparisons across cohorts and the interpretation of associations between radiomic features and disease phenotype.

The Image Biomarker Standardization Initiative (IBSI) has addressed variability across software implementations by standardizing feature definitions, processing configurations and convolutional filters [3]. Its reference data provide a common basis for evaluating numerical agreement, including the effects of preprocessing, discretization and aggregation [4], [5]. PyRadiomics and MIRP provide established CPU implementations [1], [6], but larger cohorts, more filter combinations and voxel-wise extraction can increase the computational burden enough to make large-scale quantitative imaging impractical. Voxel-wise mapping is particularly demanding when multiple texture features are calculated at each voxel within a segmented volume.

Previous work has shown that GPU acceleration can reduce this computational burden [7], [8], [9]. However, the GPU implementations of cuRadiomics, FastRad and Flash-Radiomics rely on NVIDIA hardware, whose cost may limit access for research groups with constrained budgets. Extending acceleration to integrated GPUs could broaden access without requiring a separate GPU workstation, provided that the resulting measurements retain agreement with standardized references.

Apple silicon combines high-performance computing with high-capacity, high-bandwidth unified memory and may offer a lower-cost alternative to dedicated NVIDIA GPU workstations. We therefore developed MLX-Radiomics to accelerate regional feature extraction, image filtering and voxel-wise mapping on Apple silicon (Fig. 1). We evaluated numerical agreement with IBSI references and independent calculations and compared extraction times with existing libraries. To examine agreement beyond the reference data, we compared measurements across libraries in a multimodal soft-tissue sarcoma cohort, with preprocessing applied independently under published settings.

**Fig. 1.**
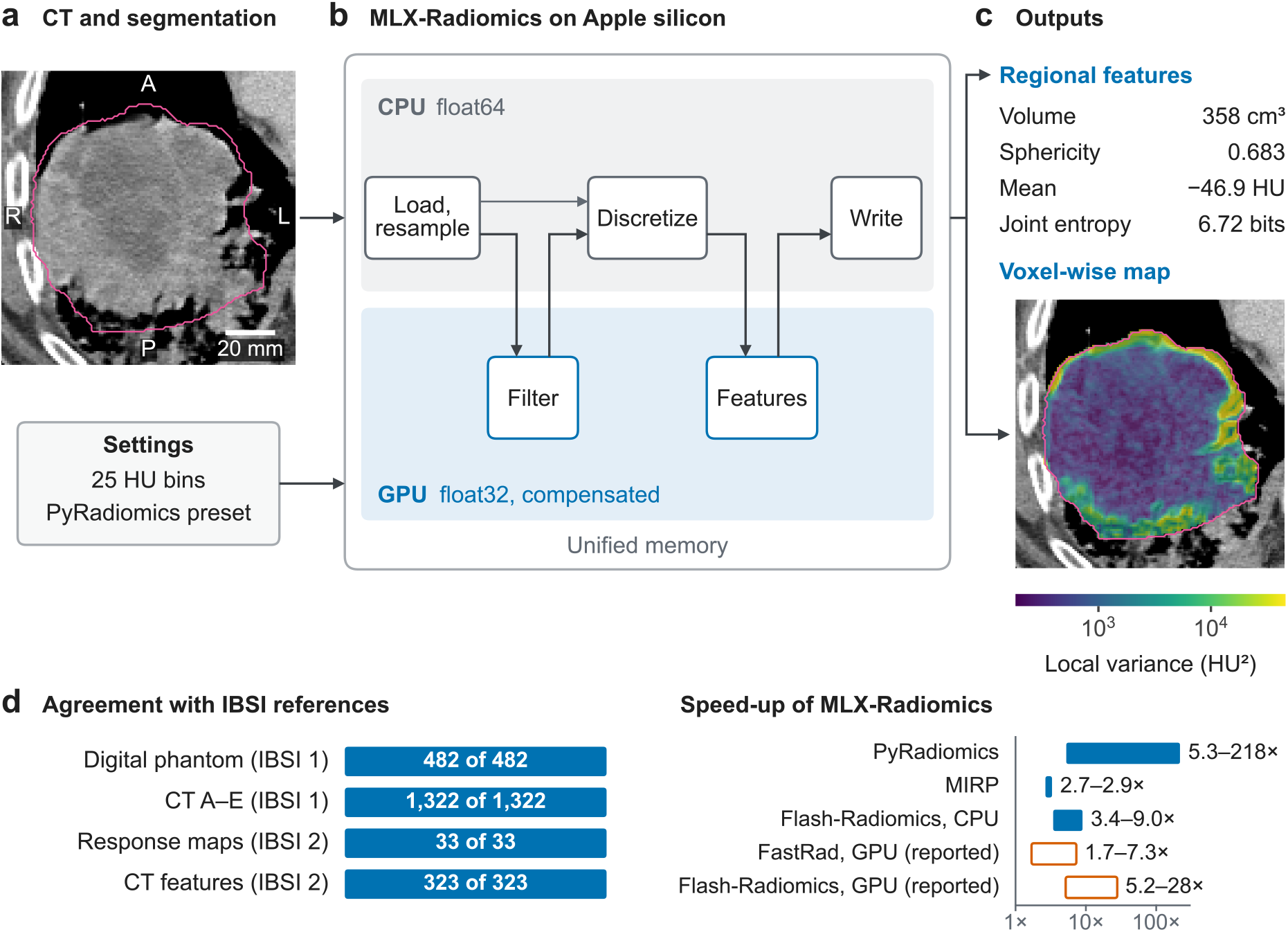
MLX-Radiomics workflow, reference agreement and computational performance. (a) CT image, segmentation (pink) and extraction settings. (b) CPU preprocessing in float64 and GPU filtering and feature calculations in float32. Both processors share unified memory; outputs are written on the CPU. (c) Regional features and a local variance map from the same CT. The map uses 3 × 3 × 3 masked neighborhoods and a logarithmic color scale with 2nd–98th-percentile limits. (d) Agreement with IBSI reference data and extraction speed-up ranges across the evaluated analyses (Fig. 3). All 2,160 reference comparisons met the specified acceptance criteria. Filled bars show measured CPU comparisons; open bars show comparisons with published GPU timings obtained on different hardware and images. Speed-up is plotted on a logarithmic axis. Image and overlay adapted from the IBSI CT.

## Feature extraction

MLX-Radiomics implements 172 IBSI feature definitions, 16 PyRadiomics-derived extensions and the IBSI 2 image filters (Fig. 1). The IBSI feature families include morphology, local intensity, intensity statistics, intensity histogram, intensity–volume histogram and texture. The texture families include gray-level co-occurrence (GLCM), run-length (GLRLM), size-zone (GLSZM), distance-zone (GLDZM), neighborhood gray-tone difference (NGTDM) and neighboring gray-level dependence (NGLDM, also called GLDM). The 16 extensions comprise three first-order statistics (total energy, entropy and uniformity), the maximal correlation coefficient and 12 two-dimensional shape measures. Image filters include mean, Laplacian of Gaussian (LoG), Laws, Gabor, separable and nonseparable wavelets, and Riesz transforms.

MLX-Radiomics uses MLX on Apple silicon, where the CPU and GPU share memory [10]. MLX supports double precision (FP64) only on the CPU [11]. Preprocessing therefore runs on the CPU in FP64, whereas filtered images and feature values are calculated on the GPU in single precision (FP32; Fig. 1b). Compensated arithmetic limits accumulation error. We evaluated the GPU results against independent calculations performed in FP64 where available. MLX-Radiomics provides ten configurable settings to handle implementation choices that IBSI leaves open and differences among software libraries (Table S1). The default settings follow IBSI reference values or recommendations in the IBSI manual. A PyRadiomics preset changes eight of these ten settings.

## Results

We evaluated MLX-Radiomics using IBSI reference images, three CT cases for timing experiments, and a cohort of 51 patients with CT, FDG PET and T1-weighted MRI (Table 1). The cohort analysis assessed feature agreement; extraction time was not measured in this cohort.

**Table 1.** Imaging data and evaluation scope. The IBSI CT was reused for reference comparisons, timing and full-region mapping. The cohort subsets comprise patients from the same 51-patient cohort.

| Dataset | Images | Image or region characteristics | Evaluation |
|---|---|---|---|
| IBSI reference images [2], [3] | Digital phantoms and reference CT | Prescribed IBSI 1 feature configurations and IBSI 2 filter configurations | Agreement with published feature values and filter response maps |
| IBSI lung CT | One CT, also used above | 204 × 201 × 60 voxels; 0.977 × 0.977 × 3 mm spacing; 125,256 segmented voxels. Map extraction was timed on a 24<sup>3</sup>-voxel crop with 12,672 segmented voxels. | Matched extraction time and memory comparisons; voxel-wise agreement over the full segmentation |
| NSCLC-Radiomics, LUNG1-001 [16] | One CT | 512 × 512 × 134 voxels; 0.977 × 0.977 × 3 mm spacing; 54,639 voxels in the GTV-1 segmentation | Regional extraction of 93 features and separate feature-class timings |
| HCC-TACE-Seg, HCC_010 [18] | One CT, cropped and tiled | Lesion trimmed to 50,613 voxels and tiled without interpolation, up to 10,122,600 segmented voxels; 0.703 × 0.703 × 2.5 mm spacing | Extraction of 93 voxel-wise maps across increasing region sizes |
| IBSI soft-tissue sarcoma cohort [13] | 51 patients; 153 images: 51 CT, 51 FDG PET and 51 T1-weighted MRI | Regional and filtered features used 1 mm isotropic spacing for CT and MRI and 3 mm for PET. Sampled maps used native spacing. | Regional and filtered feature agreement after independent preprocessing; sampled maps (six patients); resampling at image edges (eight patients) |

Dimensions and spacing describe source images unless resampling is stated. Crops, tiled volumes and repeated timing runs do not represent additional patients. Comparisons with published GPU timings used different images and hardware.

### Reference agreement and numerical accuracy

MLX-Radiomics met the comparison criteria for all 2,160 IBSI reference measurements (Fig. 1d). The Simoncelli filters required a frequency grid that included both the lowest and highest frequencies in the specified range; with the default grid, two maps and two CT values fell outside the criteria (Table S1). Two Gabor maps met the reference-comparison criteria but exceeded the stricter tolerance used for numerical validation. For these maps, independent double-precision calculations closely matched MLX-Radiomics, and both differed from the IBSI reference maps by similar amounts (Fig. 2). The LoG filters showed the same pattern, indicating that single-precision rounding did not explain the larger discrepancies with the references.

**Fig. 2.**
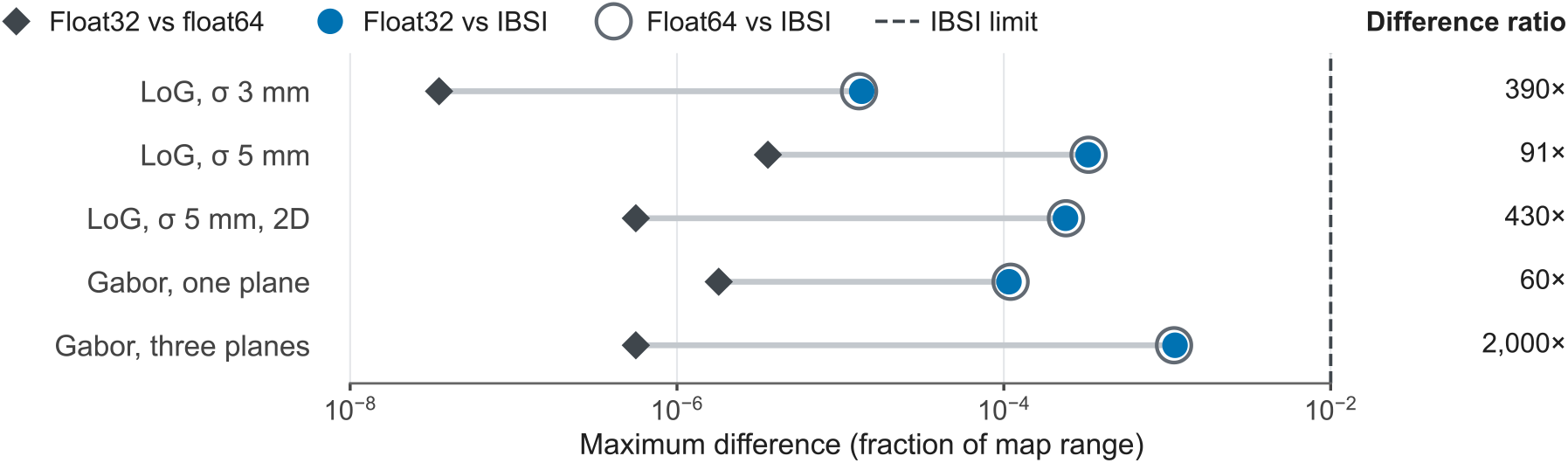
Numerical accuracy of LoG and Gabor filters. Shown are maximum absolute voxel-wise differences for three LoG and two sphere-phantom Gabor response maps, each expressed as a fraction of the corresponding IBSI map’s range. Diamonds compare MLX-Radiomics (FP32) with independent SciPy calculations (FP64). Blue dots and gray rings show differences from the IBSI reference maps for FP32 and FP64, respectively. The right column reports the maximum absolute difference between MLX-Radiomics and IBSI divided by the maximum absolute difference between MLX-Radiomics and SciPy. The dashed line marks the IBSI 2 acceptance limit of 1%.

### Feature extraction time and memory use

In the matched CT comparisons, outputs from all four analyses agreed with those from PyRadiomics or MIRP with both absolute and relative tolerances set to 10^-4^. MLX-Radiomics generated voxel-wise maps 198 to 218 times faster than PyRadiomics and extracted regional features 2.66 to 7.27 times faster than PyRadiomics or MIRP (Fig. 3a; Table S2). MLX-Radiomics also produced matching outputs 3.43 to 8.96 times faster than the fastest Flash-Radiomics CPU configuration tested for each analysis. Peak additional memory use for MLX-Radiomics remained below 600 MiB across the PyRadiomics and MIRP comparisons. However, MLX-Radiomics used more memory than PyRadiomics for regional feature extraction and LoG filtering (Fig. 3b).

**Fig. 3.**
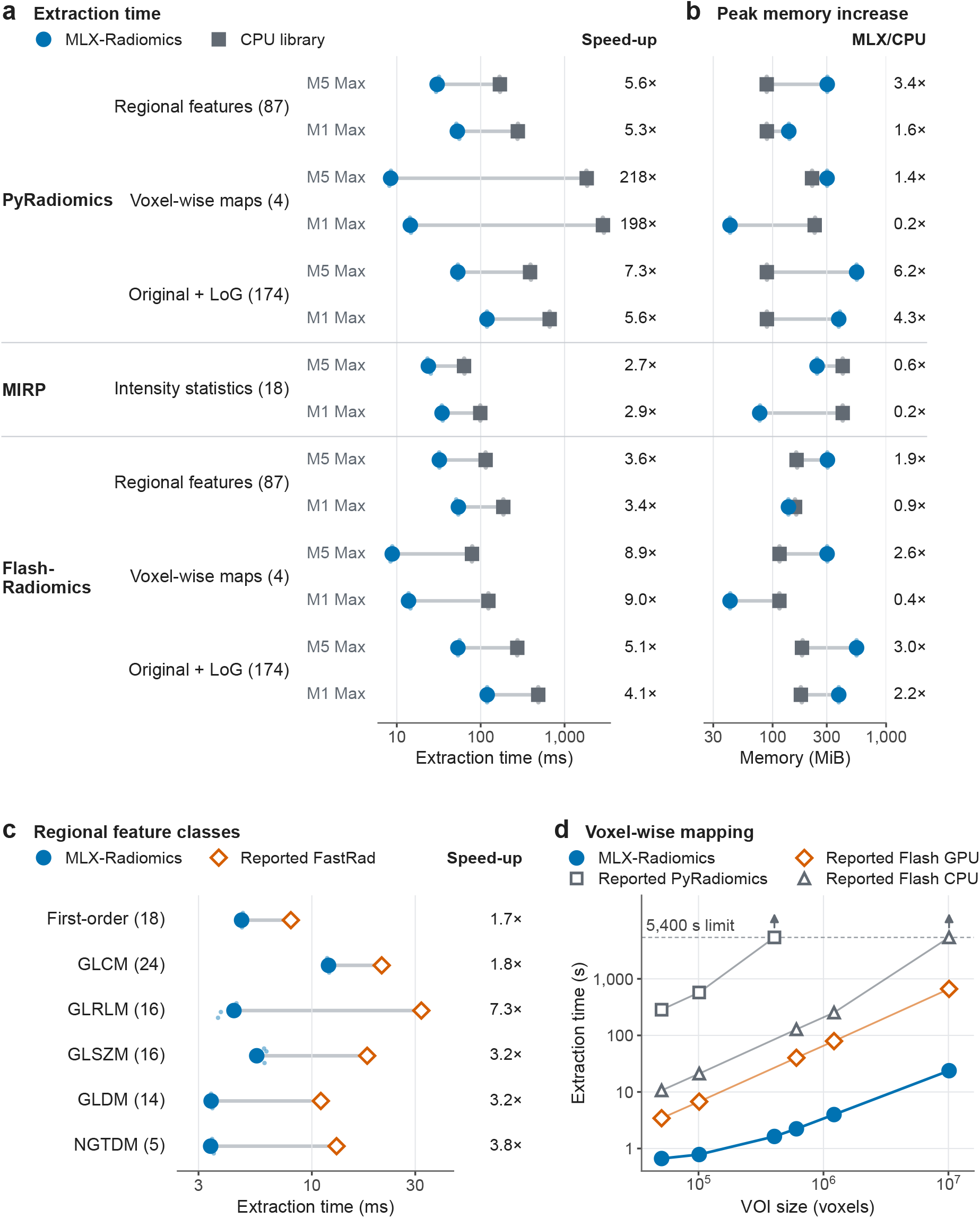
Feature extraction time and memory use. (a, b) Extraction time and peak memory increase on the IBSI CT; voxel-wise maps were computed on a 24^3^-voxel crop. MLX-Radiomics and CPU libraries were compared on each system; Flash-Radiomics used its fastest CPU setting. Large markers show the lower median of six runs; small markers show individual runs. Columns report CPU time divided by MLX time and MLX memory use divided by CPU memory use. (c) Timings by feature class for MLX-Radiomics on LUNG1-001, compared with published FastRad timings on a TCIA lung cancer CT using an RTX 4070 Ti. Each class was timed separately. (d) Extraction of 93 maps across volume-of-interest (VOI) sizes. MLX-Radiomics timings represent the median of five extractions from a tiled liver CT; published timings were obtained using tiled breast MRI (Flash GPU on an A100; Flash CPU and PyRadiomics on two AMD EPYC 7742 CPUs). In (c, d), filled markers show M5 Max measurements; open markers show published timings from different hardware and images. Arrows mark runs exceeding 5,400 s. Time and memory axes are logarithmic.

We evaluated the other libraries without modifying their source code. Constraints in their hardware support and numerical behavior limited most comparisons. Flash-Radiomics used the CPU even when configured for the Apple GPU. In that configuration, its maps agreed with MLX-Radiomics, but some regional and LoG-filtered feature values differed. FastRad’s CT feature values agreed with MLX-Radiomics, but it failed the fractional-intensity control because it uses the lower of the two middle values as the median for an even-length input. We therefore did not measure its extraction time. FastRad supported GPU extraction only through CUDA; we did not measure CUDA performance for FastRad or Flash-Radiomics.

On the LUNG1-001 CT, MLX-Radiomics extracted 93 features in 16.3 ms, compared with 721 ms for PyRadiomics. All values agreed within the specified 10^-4^ tolerances. Additional comparisons used published GPU timings obtained on different hardware and images. For individual feature classes, extraction with MLX-Radiomics was 1.7 to 7.3 times faster than reported for FastRad on a TCIA lung cancer CT using an RTX 4070 Ti [12] (Fig. 3c). For voxel-wise mapping, MLX-Radiomics generated 93 maps over 10.1 million voxels in 23.8 s, compared with 664 s reported for Flash-Radiomics on an NVIDIA A100 GPU [9]. MLX-Radiomics was faster across all tested region sizes (Fig. 3d).

### Agreement across patient images

Voxel-wise agreement with PyRadiomics was maintained across the full CT segmentation, beyond the small crop used for timing (Fig. 4a). All four maps met the agreement criteria at every voxel, with differences below 0.31% of the tolerance (Fig. 4b). We also assessed local entropy and GLCM contrast in six patients with soft-tissue sarcoma from the IBSI validation dataset [13], [14], [15]. Both features agreed with PyRadiomics at 144 sampled neighborhoods on CT, FDG PET and T1-weighted MRI under the comparison criteria described in Methods. Local entropy maps from one of these patients (STS 001) illustrate the measurements across modalities (Fig. 4c–e).

**Fig. 4.**
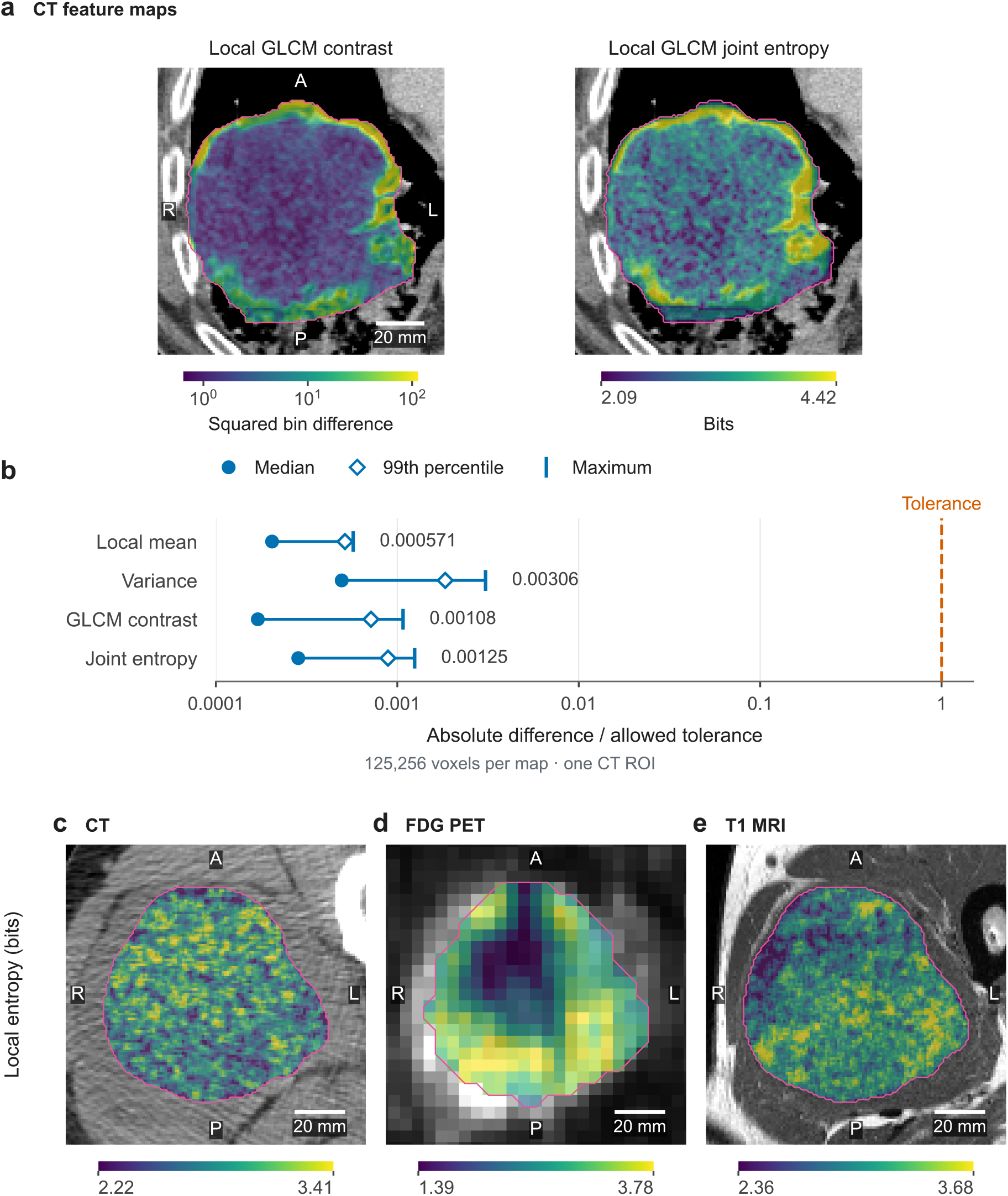
Voxel-wise radiomic feature maps. (a) GLCM contrast and joint entropy on the axial CT plane with the largest segmented cross-section, using 3 × 3 × 3 masked neighborhoods and 25 HU bins. Contrast uses a logarithmic scale; entropy is shown in bits. Color limits span the 2nd–98th percentiles. The straight lower boundary in the entropy map reflects the ROI boundary in the adjacent slice. (b) Voxel-wise agreement with PyRadiomics across 125,256 voxels per map in one CT ROI. Absolute differences are divided by the allowed tolerance, 10^-4^ + 10^-4^|PyRadiomics|. Circles, open diamonds and vertical ticks show the median, 99th percentile and maximum; labels give the maxima. Connecting lines span the median to the maximum, and the orange dashed line marks the tolerance threshold of 1 on the logarithmic axis. (c–e) Local entropy on CT, FDG PET and T1-weighted MRI from one patient with soft-tissue sarcoma, with separate color scales. Pink contours mark the regions of interest; scale bars, 20 mm. Images adapted from the IBSI CT and IBSI validation images.

We evaluated regional feature agreement across all 51 patients, each with CT, FDG PET and T1-weighted MRI (Fig. 5a). Under the published E3 settings (Table E3 of the IBSI 1 supplement [2]), MLX-

**Fig. 5.**
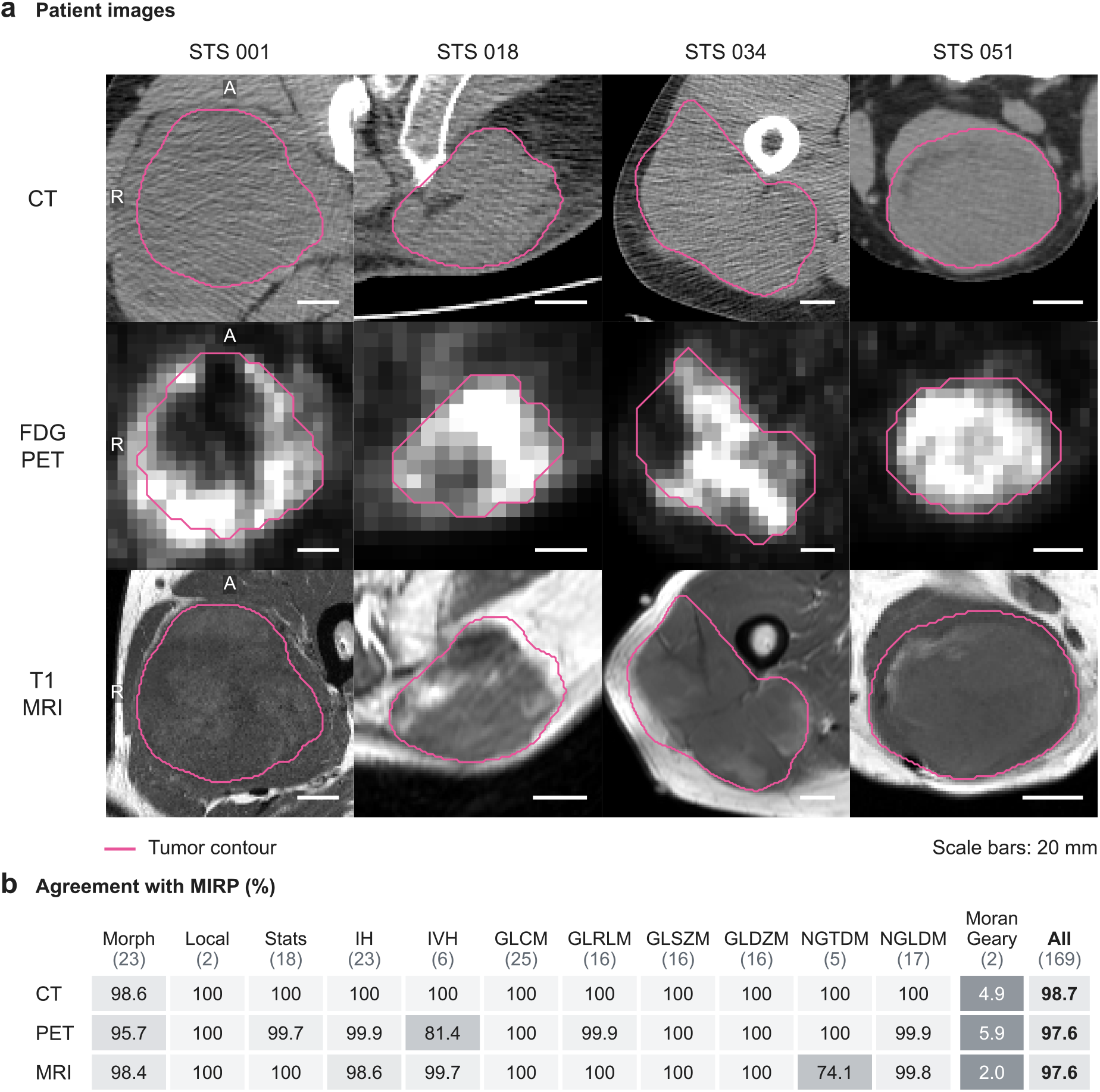
Radiomic feature agreement across imaging modalities. (a) CT, FDG PET and T1-weighted MRI from four patients in the 51-patient soft-tissue sarcoma cohort. Each column shows one patient, with pink tumor contours and 20 mm scale bars. The axial plane with the largest tumor cross-section is shown for each imaging modality. CT is displayed from −160 to 240 HU; PET and MRI use the 2nd–98th percentiles of positive intensities. R, patient right; A, anterior. Images and masks are from the IBSI validation dataset. (b) Percentage of requested values that agreed between MLX-Radiomics and MIRP across all 51 patients, grouped by imaging modality and feature type. Agreement required |MLX − MIRP| ≤ 10^-4^ + 10^-4^|MIRP|. The number of features is shown in parentheses; values that could not be calculated are counted as not agreeing. Moran’s I and Geary’s C are shown separately because MIRP estimates them using random sampling (the Monte Carlo method). Morph, morphology; Local, local intensity; Stats, intensity statistics; IH, intensity histogram; IVH, intensity–volume histogram.

Radiomics computed 25,841 of 25,857 requested values, including MRI texture features calculated by grouping image intensities into intervals 0.05 units wide, as specified. The software could not calculate 16 values of Moran’s I and Geary’s C for four CT and four MRI regions. These regions exceeded the size limit for exact calculations using every pair of voxels. MLX-Radiomics and MIRP agreed on 98.0% of requested values, with similar agreement across CT, PET and MRI (Fig. 5b). Each software package separately prepared the images and calculated the features. The comparison therefore reflected differences in both image preparation and feature calculation.

Moran’s I and Geary’s C, two measures of spatial patterns in image intensity, accounted for 277 of the 511 calculated values that differed by more than allowed across the cohort (Fig. 5b). MLX-Radiomics calculates these features exactly, whereas MIRP estimates them using random sampling (the Monte Carlo method). Excluding these two features, agreement was 99.8% for CT, 98.7% for PET and 98.8% for MRI. The remaining differences were mainly in shape measurements calculated from a model of the tumor surface (a mesh), where the median difference was 0.05%, and in NGTDM texture and intensity– volume histogram features. For these latter two groups, the libraries assign intensity levels differently.

We separately tested how image preparation affected the results in eight patients whose tumor segmentations reached the first or last slice of the image volume. When the images were placed on a new voxel grid (resampling), extending the outlined tumor region beyond these image edges produced small increases in tumor volume but substantial differences in radiomic measurements (Table 2).

**Table 2.** Effect of resampling at image edges on tumor volume and radiomic agreement. The same 24 images from eight patients were resampled using every combination of rules for extending the outlined tumor region (mask) and image intensities beyond the image edges; all other settings were identical.

| Mask extrapolation | Image extrapolation | Change in segmentation volume, %<br>Median (range) | Values outside tolerance,<br>n (% of requested) |
|---|---|---|---|
| Empty | Edge intensity repeated | 0 (reference) | 64 (1.6) |
| Empty | Mirrored | 0 (0–0) | 1,164 (28.7) |
| Edge mask copied | Edge intensity repeated | 0.91 (0.18–2.98) | 2,837 (69.9) |
| Edge mask copied | Mirrored | 0.91 (0.18–2.98) | 2,843 (70.1) |

Each condition requested 4,056 feature values (169 per image); 4,048 were computed. The number of values outside tolerance excludes the eight uncomputed values. Agreement required |MLX − MIRP| ≤ 10<sup>−4</sup> + 10<sup>−4</sup>|MIRP|. MIRP and the reference condition used empty-mask extrapolation and repeated edge intensities. Volume changes were calculated per image relative to this reference, before intensity resegmentation. Software settings are listed in Table S1.

Changing only how image intensities were assigned beyond the image edges (image extrapolation) also reduced agreement with MIRP. MLX-Radiomics and MIRP use SciPy extrapolation rules by default; PyRadiomics uses ITK (Table S1).

For the same 153 images, agreement was lower after filtering under IBSI 2 phase-3 configurations 1–7, which comprise the unfiltered image and responses to mean, LoG, Laws, Gabor and two separable wavelet filters. MLX-Radiomics computed all 18 intensity statistics for every image and filter. In both libraries, the unfiltered statistics were identical to their corresponding IBSI 1 statistics, and MLX-Radiomics agreed with MIRP on all 18 values in 152 of 153 images. After filtering, 15,200 of 16,524 values (92.0%) agreed.

## Discussion

MLX-Radiomics agreed with IBSI reference measurements and extracted features faster in the CT comparisons. The largest reductions in extraction time occurred in voxel-wise mapping, easing the computational burden of analyses of spatial variation within tumors (Fig. 4). The speed advantage was observed on both tested systems, although memory requirements were higher in some comparisons. Using the integrated GPU makes this acceleration available without a separate GPU workstation.

The numerical comparisons helped assess whether rounding during calculation explained differences from the reference measurements. Independent calculations using higher precision (double precision) closely matched the single-precision GPU results for the evaluated filters, including cases in which both differed from the IBSI reference maps. This indicates that increasing precision alone would not resolve the larger LoG and Gabor differences. For the Simoncelli filters, agreement depended on how the frequency grid was defined. These findings support single-precision calculations for the evaluated filters and highlight the need to specify the numerical rules used (Table S1).

In the patient cohort, agreement depended on both image preparation and feature calculation. At the image edge, the ITK rule for extending the tumor mask added one or two slices to the resampled tumor region. Although the resulting volume changes were small, many feature values differed. Such differences can complicate comparisons across cohorts analyzed with different software, even when feature definitions are consistent. For measures of spatial patterns in image intensity, differences arose because MLX-Radiomics calculated exact values, whereas MIRP estimated them using random sampling (the Monte Carlo method). Smaller differences remained in shape measurements based on tumor-surface models and in how intensity levels were assigned. Agreement was lower after filtering, but each library filtered the images it had prepared independently, so that comparison cannot separate the effects of filtering from those of image preparation or feature calculation. Consistent radiomic measurements therefore require agreement on how images are prepared as well as how features are calculated.

Timing experiments used three CTs. The initial extraction did not show the same speed advantage in every analysis. Extraction time was not measured in the patient cohort, so these results do not describe performance across its full range of images. The experiments compared complete software implementations and could not separate the effects of memory shared by the CPU and GPU, GPU hardware or individual software changes. The study evaluated computation and numerical agreement. It did not evaluate methods for reducing measurement differences across scanners and acquisition protocols, diagnostic performance or treatment-response prediction. The spatial maps were not compared with findings from tissue examination (histology) or clinical outcomes.

Software-specific limitations also affected the comparisons. The PyRadiomics comparison excluded six outputs because the tested MLX-Radiomics PyRadiomics preset and PyRadiomics assign different values in special cases, such as inputs with only one intensity level. The cohort comparison includes differences between libraries in image preparation, resampling and the calculation of Moran’s I and Geary’s C. Exact calculation of these two spatial measures is limited to regions of at most 1,048,576 voxels. Simoncelli configurations 8 and 9 were disabled in the MLX-Radiomics version used for the cohort filter evaluation.

MLX-Radiomics addresses two practical obstacles to quantitative imaging research: differences in measurements between software packages and the time required to calculate features. The reference comparisons and evaluation of CT, PET and MRI images showed where measurements agreed and where differences remained. Faster CT extraction on relatively inexpensive hardware reduces the computational burden of regional and voxel-wise analysis.

## Methods

We used only publicly available data, including IBSI reference data and phantoms, the IBSI validation cohort, and CT cases from The Cancer Imaging Archive (TCIA). The source datasets and their identifiers are listed in Code and data availability.

### IBSI reference comparisons and independent numerical validation

#### IBSI reference values

To determine agreement with standardized feature definitions, we evaluated the IBSI 1 digital phantom (482 comparisons) and CT configurations A–E (1,322 published values) on the GPU using the default settings. When a published tolerance was nonzero, we applied it directly. Integer-valued features had to agree within four FP32 rounding units; other values with zero published tolerance had to match after rounding both outputs and references to three significant digits, the precision of IBSI consensus submissions. For IBSI 2 response maps, every voxel had to differ from its reference by no more than 1% of the reference map’s range. IBSI 2 CT feature values were assessed against their published tolerances. The IBSI 2 reference evaluation (Fig. 1d) used MLX with a Fourier-transform correction that was absent from the build used for the cohort and timing experiments. Simoncelli and Riesz filters were disabled in those experiments. Features designated by IBSI as not standardized were excluded, as was the quartile coefficient of dispersion for IBSI 2 configuration 8.B, which lacked a consensus reference value.

#### Independent numerical calculations

The IBSI reference tolerances can exceed the differences expected from single-precision rounding. We therefore evaluated 18 mean, Laws and Gabor maps using a relative tolerance of 2 × 10^-4^ and an absolute tolerance of 2 × 10^-5^, comparing filter responses with independent FP64 calculations. Each of the 188 feature definitions and extensions was also evaluated against a case with an analytically known result or an independent FP64 calculation.

### Software comparisons

#### Runtime and agreement with CPU libraries

We required numerical agreement before comparing extraction time. Each analysis used five numerical checks covering constant intensities, non-integer intensities, irregular segmentation and IBSI reference images. The timing comparisons used the IBSI CT with 125,256 voxels in the region of interest (ROI), or a 24^3^-voxel crop for mapping. We compared MLX-Radiomics with PyRadiomics [1] for 87 intensity and texture features, four voxel-wise feature maps (local mean, variance, GLCM contrast and joint entropy), and 174 features from the original image and a 2 mm LoG response. We also compared MLX-Radiomics with MIRP [6] for 18 IBSI intensity statistics. We compared unrounded values at absolute and relative tolerances of 10^-4^ and required nonfinite values (NaN or infinity) to occur at the same locations. MLX-Radiomics used its PyRadiomics preset (Table S1) in every comparison with PyRadiomics, Flash-Radiomics and FastRad, and its defaults in comparisons with MIRP. Six outputs were excluded from the PyRadiomics comparison because their values differ in special cases under the MLX-Radiomics PyRadiomics preset evaluated in this study: skewness, kurtosis, coarseness, busyness and strength (Table S1), and the maximal correlation coefficient, which PyRadiomics sets to 1 for a single gray level.

#### Flash-Radiomics and FastRad

We compared MLX-Radiomics with Flash-Radiomics [9] and FastRad using the same images, features and numerical controls. Flash-Radiomics was tested with its CPU and Apple GPU settings; FastRad was tested on the CPU. We did not change the libraries’ feature calculations. FastRad used neighborhoods that were not restricted to the mask, so we excluded its maps from the comparison. We measured extraction time only when numerical agreement was confirmed. Initial CPU comparisons used one thread, and preprocessed images were not reused between measurements. Timing included writing all required output files. CPU libraries used their default numerical precision; MLX-Radiomics used FP32.

#### Flash-Radiomics CPU settings

We tested seven configurations on each computer, varying the number of CPU threads and the number of feature classes calculated in parallel. We used 2 to 18 threads on the M5 Max and 2 to 10 on the M1 Max. We tested both standard and OpenMP-enabled builds. Each configuration underwent the same five numerical controls and six paired timing runs. We selected the fastest configuration for each analysis from these measurements, a choice that favored Flash-Radiomics in the comparison.

#### Matched voxel-wise feature maps

To assess agreement beyond the small crop used for timing, MLX-Radiomics and PyRadiomics calculated mean, variance, GLCM contrast and joint entropy over all 125,256 voxels of the full CT ROI. We used the original voxel spacing, 25 HU bins, radius-one (3 × 3 × 3) neighborhoods restricted to the mask, and GLCM distance one. We compared unrounded values after aligning the maps by their physical coordinates, without interpolation.

#### Multimodal cohort

To assess agreement across patient images and modalities, we evaluated CT, FDG PET and T1-weighted MRI from 51 patients in the IBSI 1 soft-tissue sarcoma cohort (153 images), requesting 169 feature values per image and comparing them with independently computed MIRP results. MRI bias correction and normalization had already been applied to the distributed images. Each library independently resampled the source images to 1 mm isotropic spacing for CT and MRI and 3 mm for PET, using cubic B-spline interpolation for images and linear interpolation with a 0.5 threshold for masks. MLX-Radiomics used its default rules beyond the outermost voxel centers (Table S1). The 24 images whose masks reach the first or last slice were also evaluated with the ITK mask rule, the ITK image rule or both, with all other settings held fixed. CT values were rounded to integer HU, and the analyzed region was restricted to voxels within [−200, 200] HU; PET and MRI analyses used nonnegative intensities. Published E3 settings used averaged 3D GLCM/GLRLM features and MRI bin widths of 0.05 for texture/histogram features and 0.01 for intensity–volume histograms. Finite values agreed if |MLX − MIRP| ≤ 10^-4^ + 10^-4^|MIRP|. Agreement percentages included all requested values in the denominator, including values that could not be calculated. MLX-Radiomics calculated Moran’s I and Geary’s C using exact weights for every pair of voxels, whereas MIRP used a reproducible random-sampling estimate (deterministic Monte Carlo approximation). For IBSI 2, each library filtered its own preprocessed images with phase-3 configurations 1–7, using mirror padding, and the 18 intensity statistics were compared under the same tolerance.

### Voxel-wise feature maps and image display

The color scale for each feature map spanned the 2nd–98th percentiles of values within the region. Variance and GLCM contrast used logarithmic scales. CT window level and width were 40 and 400 HU, respectively. Images retained native spacing and were displayed using nearest-neighbor interpolation; overlays used 85% opacity. The CT and variance map in Fig. 1 share the same physical crop. Multimodal maps (Fig. 4c–e) used 3 × 3 × 3 masked neighborhoods at native spacing; CT intensities were restricted to −200 to 200 HU, and PET and MRI intensities to nonnegative values. Bin widths were 10 HU for CT, 0.25 SUV for PET and 10 stored units for MRI. The MRI bin width was chosen for illustration and differs from the published 0.05-unit width used for cohort feature extraction. For six patients, eight neighborhoods per image (144 in total) were compared with PyRadiomics; all comparisons met absolute and relative tolerances of 10^-5^ and 10^-4^, respectively.

### Feature extraction time and memory use

The four PyRadiomics and MIRP analyses and the Flash-Radiomics and FastRad comparisons were performed on a MacBook Pro with an Apple M5 Max (18-core CPU, 40-core GPU, 128 GiB unified memory) and a system with an Apple M1 Max (10-core CPU, 32-core GPU, 64 GiB unified memory). Both systems used AC power and ran macOS and Python with the same software, images and feature selections. All other timing experiments used the M5 Max. Extraction time and memory use were measured with Perfkit. Desktop applications and background services remained running.

For Table S2, Fig. 3a–b and the Flash-Radiomics and FastRad comparisons, timing covered the full pipeline, from image loading to output writing. We made six paired comparisons, alternating the order of the CPU library and MLX-Radiomics. Each run included an initial extraction followed by three repetitions. We report the median of the three repetitions within each run and, across the six run medians, the lower of the two middle values (the lower median). The initial extraction, which includes setup, is reported separately. Speed-up is the CPU library’s extraction time divided by MLX-Radiomics extraction time. The 96.875% paired-sign confidence intervals describe variation across the six runs. These were repeated measurements of one CT, not observations from different patients. Memory use is the peak increase above the memory used before extraction, in MiB (2^20^ bytes).

The LUNG1-001 CT contained 512 × 512 × 134 voxels at 0.977 × 0.977 × 3 mm, with 54,639 voxels in the GTV-1 segmentation. Timing began with images and masks already in memory. We used the original voxel spacing, 25 HU bins and distance one. All 93 outputs met absolute and relative tolerances of 10^-4^. Each implementation was tested six times, with seven repeated extractions in each run. We report the lower median of the six run medians. Feature classes were also measured separately; these times include repeated image preparation and therefore do not sum to the time for all 93 features.

For the map-size comparison, we tiled the HCC_010 liver-lesion image and mask, trimmed to 50,613 voxels, to produce larger volumes without interpolation. We used 25-unit bins and radius-one masked neighborhoods. We timed five repeated extractions at each volume size. Timing included preprocessing, feature calculation and compressed NIfTI output. FastRad and Flash-Radiomics comparison times came from the published reports and used the authors’ hardware and images. The Flash-Radiomics report does not specify the bin width used for its tiled breast MRI.

Separate GPU profiling on the M5 Max confirmed GPU use in all four analyses. Profiling was disabled during timing measurements.

## Data Availability

All source imaging data are publicly available from the IBSI and TCIA repositories linked below. Results are reported in the manuscript and supplementary tables.

https://github.com/theibsi/data_sets/tree/6da96021bc91faf4c0cb7fd7fa56a4225d2064a8

https://github.com/theibsi/ibsi_2_reference_data/tree/5404579fb3e0d17e8db421f0e82d64ce2432ec03

https://www.cancerimagingarchive.net/collection/soft-tissue-sarcoma/

https://www.cancerimagingarchive.net/collection/nsclc-radiomics/

https://www.cancerimagingarchive.net/collection/hcc-tace-seg/

## Author contributions

Joseph Sandoval and Alfredo Sandoval Jr. contributed equally to software implementation, experiments and benchmarking, comparisons with reference implementations, data analysis and interpretation, and figure preparation. Alfredo Sandoval Jr. directed the scientific work, including the research questions, methods and evaluation design, and wrote the manuscript. Joseph Sandoval edited the manuscript. Alfredo Sandoval Jr. and Arsalan Saleem supervised the project. All authors approved the final manuscript.

## Code and data availability

MLX-Radiomics is maintained at https://github.com/MedMLX/mlx-radiomics.

All data used in this study are publicly available; source patient image volumes are not redistributed. The IBSI 1 and IBSI 2 digital phantoms (CC BY 4.0) and the IBSI lung cancer CT phantom (CC BY-NC 3.0; doi:10.17195/candat.2016.08.1) were obtained from the IBSI data sets repository. The IBSI validation cohort (CC BY 3.0) is derived from the TCIA Soft-tissue-Sarcoma collection [13].

The LUNG1-001 and map-scaling benchmarks (Fig. 3c–d) used public CT cases from the TCIA NSCLC-Radiomics [16], [17] and HCC-TACE-Seg [18] collections. IBSI 2 reference response maps and feature values (CC0 1.0) were obtained from ibsi_2_reference_data.

## Ethics

This study used only publicly available, de-identified imaging data and published analysis data.

## Use of AI tools

Codex (OpenAI) was used to assist with source-code analysis and manuscript preparation. The authors reviewed and edited the text and take full responsibility for its content.

