## Supplementary Tables S1 and S2 for "MLX-Radiomics: accelerating standardized regional and voxel-wise radiomics for CT, PET and MRI"

**Table S1.** Numerical and preprocessing conventions in MLX-Radiomics.

| Measurement or convention | MLX-Radiomics default | MLX-Radiomics PyRadiomics preset |
|---|---|---|
| Principal axes for shape features[a] | Sample covariance, divisor $N - 1$ | Divisor N |
| Ellipsoid surface area for approximate enclosing ellipsoid area density[b] | Series printed in the IBSI manual | Exact area |
| Kurtosis[c] | Excess kurtosis | Without subtracting 3 |
| Skewness and kurtosis of constant intensities[d] | Zero | Not a number (NaN) |
| NGTDM singular cases[e] | Coarseness capped at $10^6$; busyness zero with one occupied gray level; strength zero when differences sum to zero | Unadjusted formulas |
| NGTDM and NGLDM neighborhoods[f] | Closed radius | Configured distance list |
| Fixed-bin-size origin without an explicit anchor[g] | Lower resegmentation bound, or the ROI minimum if no bound is specified | Multiples of the bin width |
| Gray level of constant intensities with a fixed bin count[h] | First bin | Middle bin |
| Mask extrapolation beyond the outermost voxel centers during resampling[i] | Empty (SciPy) | Edge mask copied (ITK) |
| Image extrapolation beyond the outermost voxel centers during B-spline resampling[j] | Edge intensity repeated (SciPy; IBSI manual) | Mirrored (ITK) |
| Number of gray levels in normalized inverse difference and moment | Number of discretization bins | – |
| Histogram mode ties | Bin closest to the mean, then the lower bin | – |
| Simoncelli frequency grid | DFT grid (manual text); endpoint-inclusive grid selectable, matching the reference maps | – |

Both columns describe the MLX-Radiomics implementation evaluated in this study. PyRadiomics itself uses the default conventions listed here for constant-intensity moments and NGTDM singular cases; the evaluated MLX-Radiomics preset does not. A dash indicates no change with the preset. The final three conventions are fixed or selected per filter.
Software settings: [a]`covariance_divisor`; [b]`ellipsoid_area`; [c]`kurtosis`; [d]`zero_variance_moments`; [e]`ngtdm_singular_cases`; [f]`neighborhood_support`; [g]`bin_origin`; [h]`constant_roi_bin`; [i]`mask_extrapolation`; [j]`spline_boundary`.

**Table S2.** Radiomic feature extraction times.

| Analysis<br>Comparator | Values compared | Subsequent extractions, ms | | Speed-up (CI) | Initial extraction, ms | |
|---|---|---|---|---|---|---|
| | | Comparator | MLX-Radiomics | | Comparator | MLX-Radiomics |
| **M5 Max** | | | | | | |
| Regional features (87)<br>PyRadiomics | 435 | 170 | 30.3 | 5.60×<br>(5.14–5.81) | 203 | 225 |
| Voxel-wise maps (4)<br>PyRadiomics | 61,664 | 1,837 | 8.44 | 218×<br>(199–228) | 1,903 | 147 |
| Unfiltered and LoG (174)<br>PyRadiomics | 870 | 388 | 53.4 | 7.27×<br>(7.27–7.43) | 423 | 270 |
| Intensity statistics (18)<br>MIRP | 90 | 63.3 | 23.8 | 2.66×<br>(2.52–3.00) | 1,192 | 169 |
| **M1 Max** | | | | | | |
| Regional features (87)<br>PyRadiomics | 435 | 277 | 52.7 | 5.26×<br>(4.94–5.42) | 328 | 267 |
| Voxel-wise maps (4)<br>PyRadiomics | 61,664 | 2,868 | 14.5 | 198×<br>(183–203) | 2,938 | 166 |
| Unfiltered and LoG (174)<br>PyRadiomics | 870 | 664 | 119 | 5.59×<br>(5.51–5.60) | 712 | 346 |
| Intensity statistics (18)<br>MIRP | 90 | 98.9 | 34.6 | 2.86×<br>(2.67–2.88) | 1,874 | 182 |

Times are the lower median of six paired runs on one CT per computer. The comparator used the CPU and MLX-Radiomics used the GPU. Initial extraction times include setup; subsequent times exclude it. Speed-up is the comparator time divided by MLX-Radiomics time, with 96.875% confidence intervals across the six paired runs. Voxel-wise maps used a $24^3$-voxel crop; LoG used a scale of 2 mm.

Values compared are the outputs from five numerical controls per analysis, separate from the timing runs. All compared values met the $10^{-4}$ absolute and relative tolerances. CI, confidence interval; LoG, Laplacian of Gaussian.
